# Systematic Review and Meta-analysis of the Effectiveness and Safety of Hydroxychloroquine in COVID-19

**DOI:** 10.1101/2020.05.07.20093831

**Authors:** Tzu-Han Yang, Chian-Ying Chou, Yi-Fan Yang, Yi-Ping Yang, Chian-Shiu Chien, Aliaksandr A. Yarmishyn, Tzu-Ying Yang, Shih-Hwa Chiou, Yuh-Lih Chang

## Abstract

**Backgrounds:** Since COVID-19 outbreak, various agents have been tested but no proven effective therapies have been identified. This has led to a lot of controversies among associated researches. Hence, in order to address the issue of using hydroxychloroquine (HCQ) in treating COVID-19 patients, we conducted a systematic review and meta-analysis.

**Methods:** A thorough search was carried out to find relevant studies in MEDLINE, medRxiv, PubMed, Cochrane Database, China Academic Journals Full-text Database and Web of Science. Two investigators independently reviewed 274 abstracts and 23 articles. The trials which evaluated HCQ for treatment of COVID-19 were included for this systematic review. Two investigators assessed quality of the studies and data extraction was done by one reviewer and cross checked by the other.

**Results:** Five trials involving 677 patients were included while conducting the meta-analysis. Compared with the control group, HCQ with or without azithromycin (AZI) showed benefits in viral clearance of SARS-CoV-2 (odds ratio (OR) 1.95, 95% CI 0.19-19.73) and a reduction in progression rate (OR 0.89, 95% CI 0.58-1.37), but without demonstrating any statistical significance. This systematic review has also suggested a possible synergistic effect of the combination therapy which included HCQ and AZI. However, the use of HCQ was associated with increased mortality in COVID-19 patients.

**Conclusions:** The use of HCQ with or without AZI for treatment of COVID-19 patients, seems to be effective. The combination of HCQ and AZI has shown synergistic effects. However, mortality rate was increased when the treatment was conducted with HCQ.

**Summary of the article’s main point:** This study demonstrates that the use of hydroxychloroquine with or without azithromycin might have benefits in viral clearance of SARS-CoV-2 and reduction of progression rate, but was associated with increased mortality in COVID-19.

## INTRODUCTION

In December 2019, the sudden outbreak of newly detected coronavirus (SARS-CoV-2) resulted in spread of coronavirus disease (COVID-19) to other countries, leading to a global pandemic. This virus was initially reported in the Hubei province, Wuhan, China [1]. As of April 27, 2020, more than 3 million people have been rapidly affected by COVID-19 worldwide and over two hundred thousand people have died in more than 200 countries due to this deadly disease [2]. As COVID-19 is spreading at a fast pace therefore without wasting time researchers are working tirelessly to find an effective treatment for this disease. However, despite all the efforts, there is no proven, effective, pharmacological treatment available currently except following infection control and preventive strategies such as personal protective measures and social distancing.

Various active clinical trials are underway to detect effectiveness of ribavirin, remdesivir, lopinavir/ritonavir and hydroxychloroquine (HCQ) for COVID-19 treatment. The pathogenic mechanisms of SARS-CoV-2 might be related to its impact on immune system since SARS-CoV-2 can infect T cells [3]. Therefore, recently conducted studies have drawn a lot of attention to the possible benefit of HCQ, an antimalarial agent with an immunomodulatory activity which has been used at a large-scale for the treatment of rheumatic disease mainly because of its relatively low price and considerable safety.

Several studies have speculated that HCQ has a potent antiviral activity against SARS-CoV-2 and might be the choice in treating COVID-19 patients [4–6]. However, *in vitro* studies may not complement to clinical studies and a lot of controversial findings are noticed among these researches [7–16]. Moreover, most of the published studies had small sample size [7–9, 13] or conducted a non-randomized clinical trial (non-RCT) [8, 10–12, 14, 15], which are major limitations in deriving a sound outcome.

Many review articles which have discussed the use of HCQ in COVID-19 patients showed limitation as they included only few relevant studies, significant Chinese studies were omitted and analysis of treatment effects was not quantified while formulating the results [17–19]. Recently, two systematic review with meta-analysis were published which had small sample size and there was no clear clarity with regard to the effects of using HCQ alone, or when combined with azithromycin (AZI), for the treatment of COVID-19 [20, 21]. These systematic reviews did not include any national trials and population of non-Hispanic black while conducting their research [20, 21].

On April 24, FDA issued a warning against use of HCQ and chloroquine for treating patients with COVID-19 due to its serious adverse effect, such as heart arrhythmias being reported in some patients [22]. Therefore, efficacy and safety of HCQ for patient with COVID-19 is still uncertain. In view of severe outbreak of COVID-19 and rapid increase of studies being conducted, it is crucial to systematically evaluate the relevant studies in a rigorous manner. Hence, we conducted a systematic review and meta-analysis to demonstrate the significance of present evidence regarding benefits and safety of HCQ use, for the treatment of COVID-19 patients.

## METHODS

### Data search

We systematically searched the MEDLINE, medRxiv, PubMed, China Academic Journals Full-text Database, Cochrane database and Web of Science database up till April 27, 2020, with no language restrictions taken into consideration. In addition, relevant articles were also obtained from the reference list of reports which were identified using this search strategy. The main key words included the following terms, “COVID-19”, “SARS-CoV-2” and “hydroxychloroquine”.

### Study selection

Two reviewers reviewed 274 abstracts and 23 full-text articles independently. Our study included only comparative studies that examined the efficacy or safety of HCQ with or without AZI in comparison with standard treatment based on factors such as virological cure, degree of progression to severe illness and all-cause mortality as identified in COVID-19 patients. Out of all the selected search material, 257 articles were excluded due to their inappropriate study design (review, editorial News and letter). Among these, we also excluded a RCT as the results showed that 8 % (n=6) of the patients assigned to HCQ group did not receive any dose of HCQ and this might have caused misclassification bias in the study [23].

### Quality assessment

One reviewer extracted study-level data into standardized working tables, and the other checked data accuracy. Two independent reviewers critically appraised the eligible articles by using the Modified Downs and Black risk assessment scale [24]. This scale consists of 27 items which assesses different study characteristics such as internal validity, statistical power and external validity. Publication bias was also assessed by using a funnel plot.

### Data synthesis and statistical analysis

The efficacy outcome of interest in this article was viral clearance and progression of disease. The safety outcome which we planned to investigate through this review, was mortality. We analyzed the main beneficial outcome based on the consistency and availability of results which were reported across various recruited trials such as the level of viral clearance noticed after the treatment, which was defined as a negative result of SARS-COV-2 after detecting it by real-time reverse transcription-PCR. We performed the meta-analysis by using Mantel-Haenszel method for dichotomous data. In our present study, either fixed-effect model or random effect model was used according to the level of heterogeneity. The odds ratio (OR) was used as the common measure of association across studies. We assessed statistical heterogeneity through the *I*^2^ index. Meta-analysis was undertaken by using Cochrane Collaboration Review Manager (RevMan) software version 5.3.

### Subgroup analysis

Subgroup analysis was carried out by including the symptomatic clinical status and age (≥18 years old) so as to assess the impact of these variables on the outcome.

## RESULTS

### Study characteristics

There was a total of 677 participants, and each trial included in this research had sample size ranging from 30 to 368 [7–11]. Supplementary Table 1 shows the main characteristics of the eligible studies included in the research. Follow-up duration of included trials ranged from 6 to 7 days. Among these five included trials, four trials mentioned their exclusion criteria as - the patients who had known allergy to HCQ or any other known contraindication in terms of usage of the study drugs, such as retinopathy or pregnancy [7–9, 11]. However, one trial was conducted without considering any exclusion criteria and where the average age of participants was reported to be 62.6 years [10].

### Quality of included studies

The risk of bias assessment is reported in Supplementary Figure 1. The average Downs and Black score was 19, with a range between 18 and 22 (A higher score indicates less bias). Various studies have shown different degree of bias wherein the RCT conducted by Chen et al has displayed the lowest risk of bias [9] and the open-label non-RCT had shown the highest risk of bias [8]. One Chinese trial was described as randomized, although the method of randomization, allocation concealment and blinding of assessors during outcome evaluation was not mentioned [7] whereas another randomized control trial properly described the method of randomization and double-blinding [9]. With regard to the study design, one French trial was designed as open-label non-randomized trial [8] and two other studies were planned as controlled retrospective studies [10, 11]. A trial reported a loss of follow-up by participants while undergoing treatment hence they were excluded during analysis planning due to their withdrawal from the study [8].

### Effect on viral clearance

Two trials were included for meta-analysis, assessing a total of 66 participants [7, 8]. We found no significant evidence to verify whether HCQ with or without AZII used for the treatment was effective in reducing viral carriage (OR 1.95, 95% CI 0.19-19.73). There was substantial and statistically significant heterogeneity present while conducting this analysis (*I*^2^ = 62%) (Figure 2A). As for the treatment undertaken by using HCQ alone, the meta-analysis showed an increment by 1.74 times as compared with control group (95% CI 0.51-5.91, *I*^2^ = 35%) while evaluating the viral clearance (Figure 2B).

**Figure 1.**
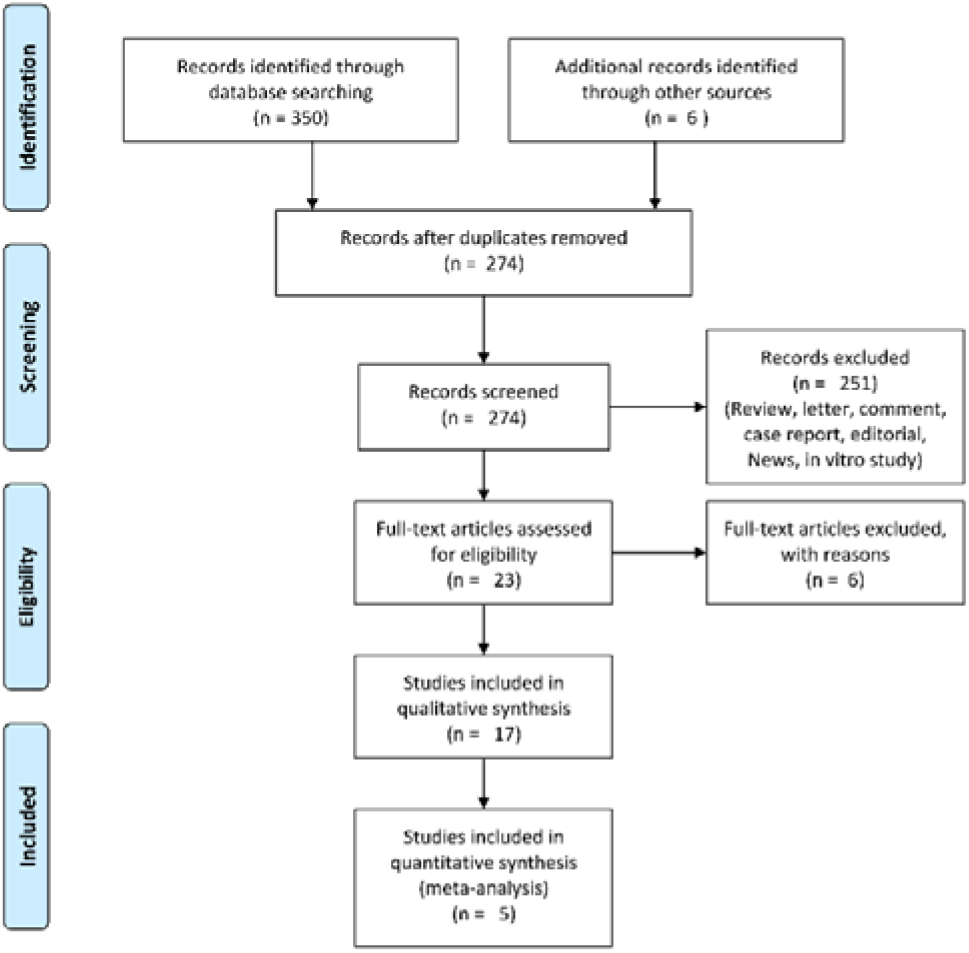
Prisma flow chart of included studies.

**Figure 2.**
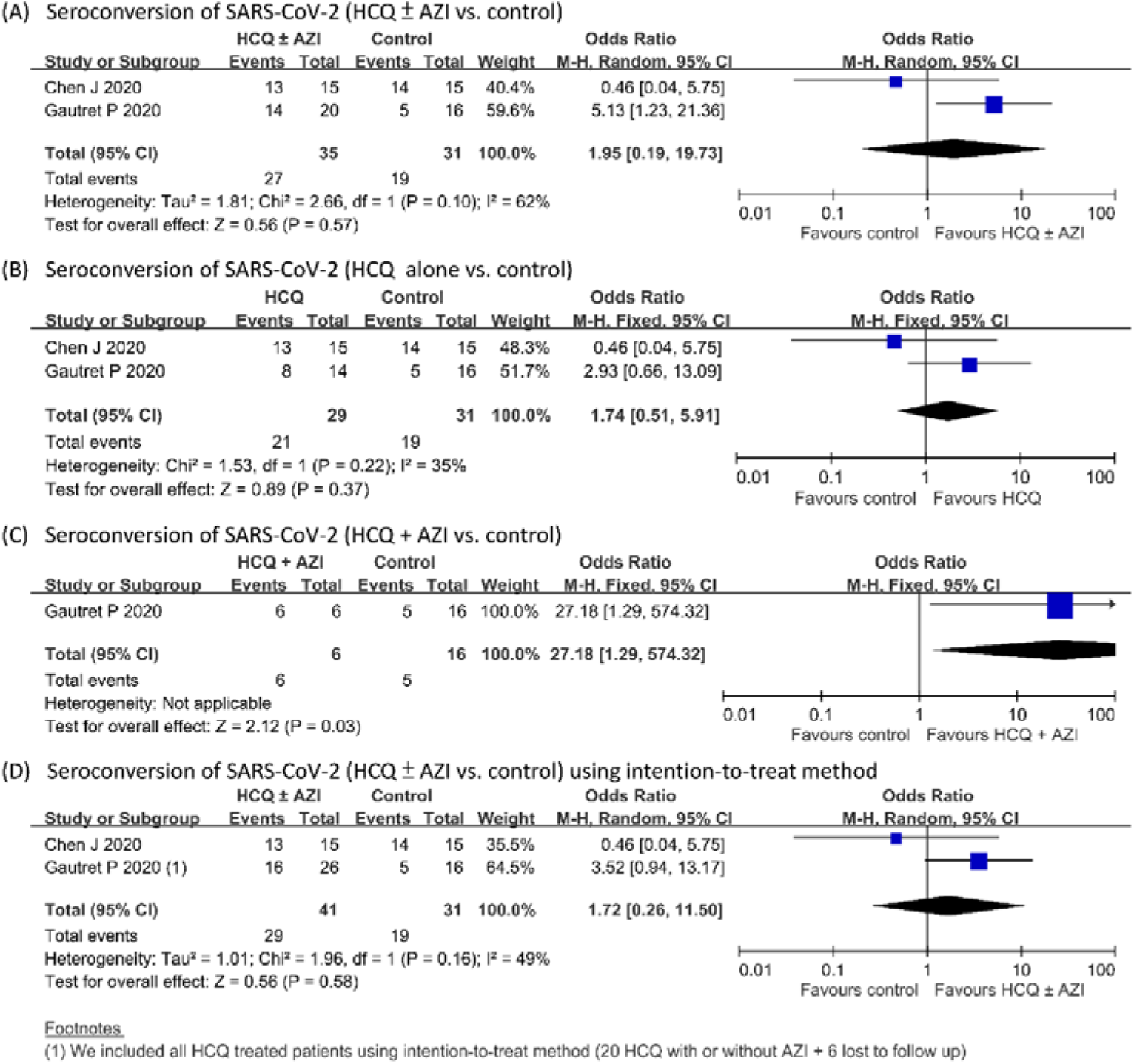
Viral clearance of SARS-CoV-2 in COVID-19 patients (Hydroxychloroquine-based treatment versus control). HCQ ± AZI: the use of hydroxychloroquine with or without azithromycin; HCQ alone: the use of hydroxychloroquine without azithromycin; HCQ + AZI: combination of hydroxychloroquine and azithromycin; CI: confidence interval.

Figure 2C showed the comparing effectiveness of combination therapy of HCQ and AZI with control group, and the results indicated that there was a significant reduction in viral load when combination therapy was used (OR 27.18, 95% CI 1.29-574.32). Figure 2D showed pooled analysis which implied a trend that treatment by HCQ with or without AZI was effective in reducing viral load in Covid-19 patients (OR 1.72, 95% CI 0.26-11.50, *I*^2^ = 49%).

### Antiviral effect of HCQ monotherapy or AZI-based combination therapy

We conducted another meta-analysis to compare the antiviral effects of using HCQ alone, or when combined with AZI and the results showed that there is an inclination of more benefits being detected with regard to reduction in viral load in the combination group (OR 9.94, 95% CI 0.47-210.41) (Figure 3).

**Figure 3.**
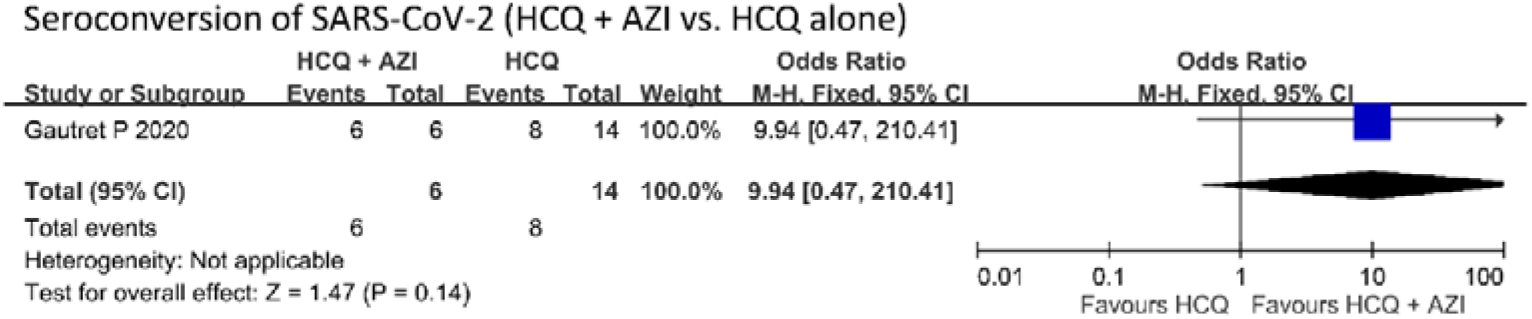
Viral clearance of SARS-CoV-2 in COVID-19 patients (Hydroxychloroquine plus azithromycin versus hydroxychloroquine alone). HCQ + AZI: combination of hydroxychloroquine and azithromycin; HCQ alone: the use of hydroxychloroquine without azithromycin; CI: confidence interval.

### Progression to severe illness

During the research period, progression to severe illness as seen in COVID-19 patients in association with the study drug was noticed in all the five included trials [7–11]. Although, quantitatively lower rate of progression to severe illness was reported in the HCQ usage groups, whether with or without AZI combination, but there was no significant difference documented when compared with control groups (OR 0.89, 95% CI 0.58-1.37, *I*^2^ =30%) (Figure 4A). We analyzed the efficacy of treatment in association with worsening of disease and the research findings showed a more favorable outcome in the study drug groups. (Figures 4B and 4C).

**Figure 4.**
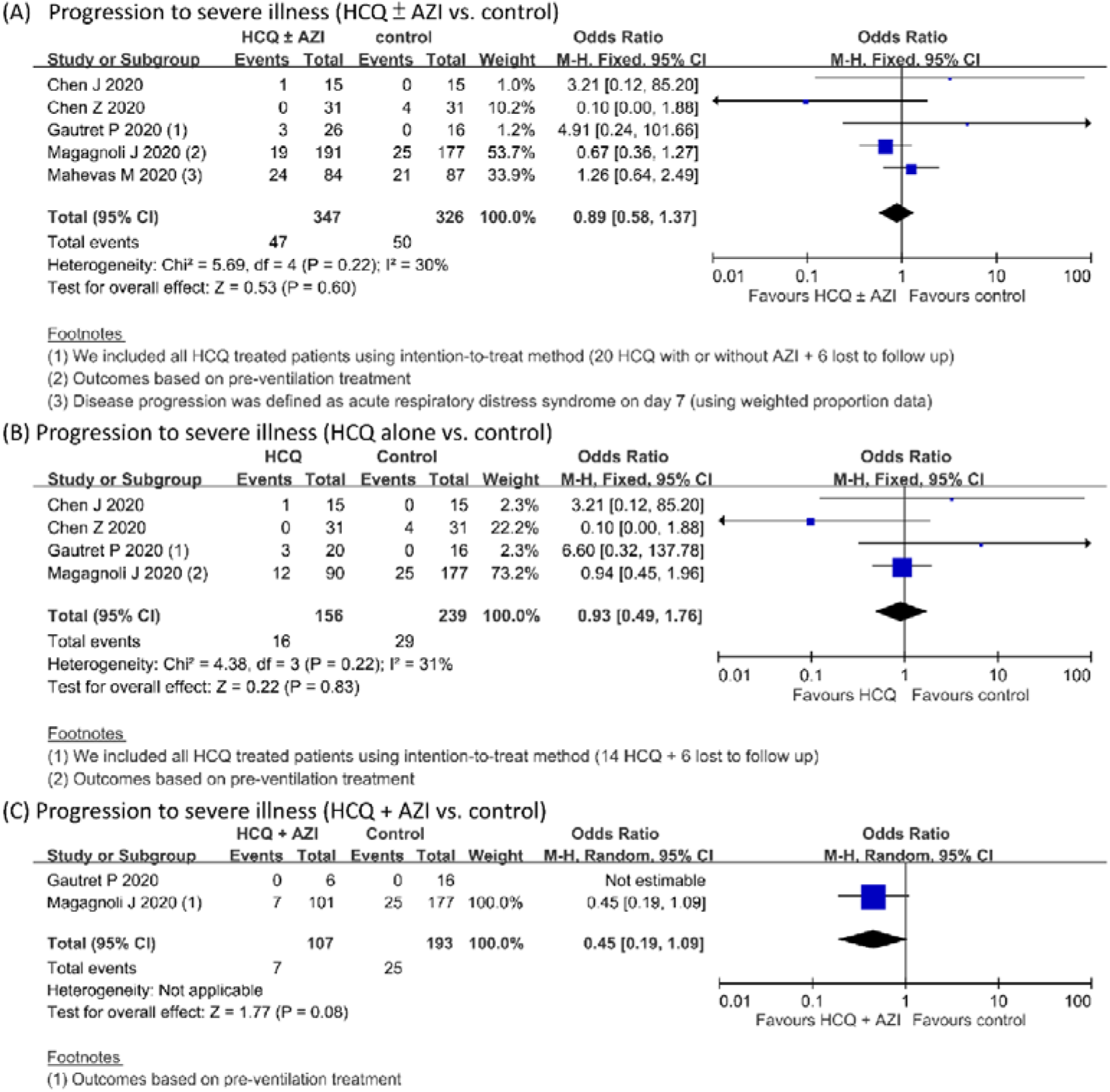
Progression to severe illness in COVID-19 patients. HCQ ± AZI: the use of hydroxychloroquine with or without azithromycin; HCQ alone: the use of hydroxychloroquine without azithromycin; HCQ + AZI: combination of hydroxychloroquine and azithromycin; CI: confidence interval.

### All-cause mortality

For analyzing all-cause mortality, we included three trials which had relevant results associated with regard to mortality [7, 8, 10]. The research findings concluded that HCQ use, whether in combination with AZI or not, was associated with increased mortality in COVID-19 patients (Figures 5A-C)

**Figure 5.**
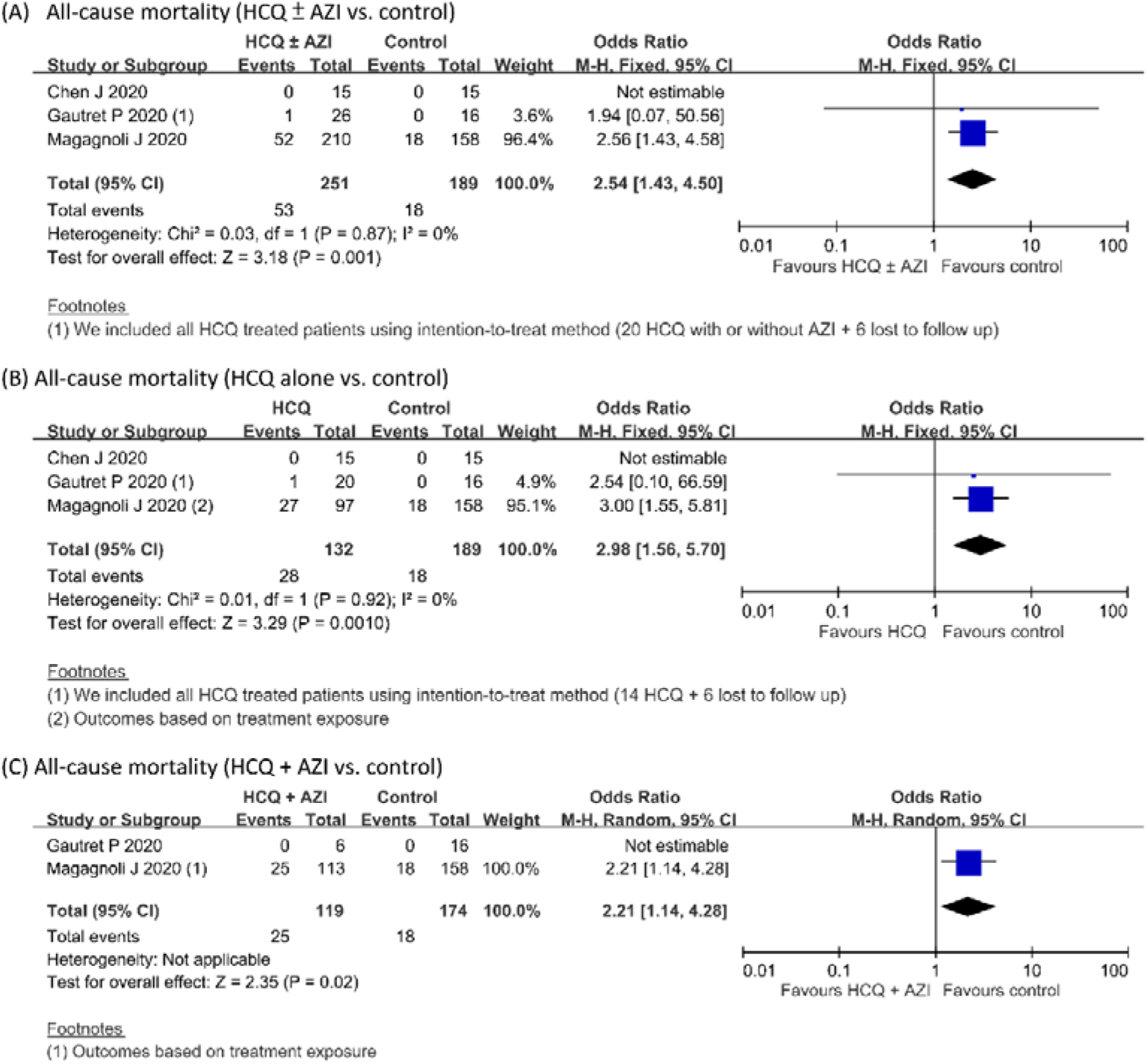
All-cause mortality in COVID-19 patients. HCQ ± AZI: the use of hydroxychloroquine with or without azithromycin; HCQ alone: the use of hydroxychloroquine without azithromycin; HCQ + AZI: combination of hydroxychloroquine and azithromycin; CI: confidence interval.

### Death or progression to severe illness

The overall pooled OR was 1.00 for death or progression to severe illness in patients treated with HCQ, with or without AZI versus control groups for four studies that had complete data regarding the same [7–9, 11] (95% CI 0.27-3.75, *I^2^* = 33%) (Figure 6A). However, the pooled OR was 1.37 when a comparison of HCQ alone versus the control group was conducted (95% CI 0.09-21.97, *I^2^* = 59%), presented in Figure 6B.

**Figure 6.**
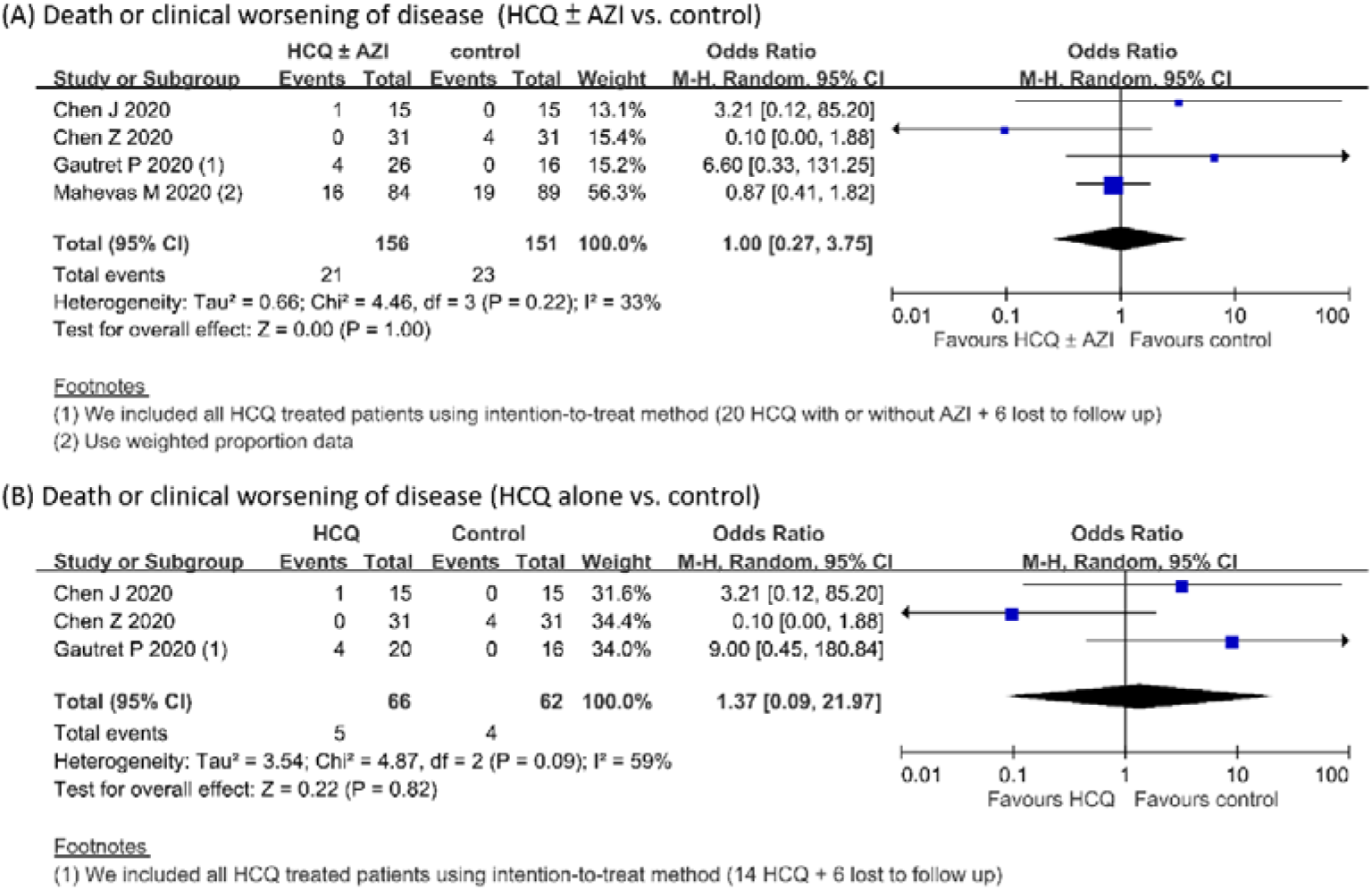
Death or clinical worsening of disease in COVID-19 patients. HCQ ± AZI: the use of hydroxychloroquine with or without azithromycin; HCQ alone: the use of hydroxychloroquine without azithromycin. CI: confidence interval.

### Subgroups analysis

Our data demonstrated that variables of symptomatic clinical status (e.g. upper respiratory tract infection or lower respiratory tract infection symptoms) as well as age did not possess significant modulation effect in viral clearance under the use of HCQ alone or in combination with AZI (Supplementary Figures 2 and 3).

### Publication bias

In total, five trials were included in the present study and were appropriately evaluated for publication bias by conducting funnel plot analysis. As shown in Supplementary Figure 4, there was no obvious publication bias reported.

## DISCUSSION

To solve the COVID-19 pandemic, it is urgently required to find effective pharmacological agents. HCQ is a repositioning candidate, still with reports of unclear efficacy and safety issue when used for the treatment of COVID-19. Therefore, we conducted a systematic review and meta-analysis to address not only the efficacy and safety issue of HCQ, but also to explore its efficacy when used in combination with AZI for treating COVID-19 patients. The present study yielded several important findings. Firstly, HCQ might show benefits in viral clearance of SARS-CoV-2 and a reduction in the risk of disease progression, although no statistically significant findings were reported. Secondly, the combination therapy of HCQ and AZI seems to show a synergistic effect. Thirdly, use of HCQ was associated with an increased mortality. To the best of our knowledge, this study is the first systematic review and meta-analysis which has addressed the issue of combining HCQ and AZI in COVID-19 patients, by summarizing available results extracted from clinical trials.

Based on only two trials that were limited in small sample size (66 patients), short observation period (6-7 days) and short duration of drug treatment (5-10 days) [7, 8], we revealed a trend of benefits in viral clearance of SARS-CoV-2 in HCQ monotherapy or combined with AZI. Although subsequent analysis for symptomatic and adult subpopulations did not reach statistically significant in our study, several recent published case reports might coordinate to our suggestions in present study. Gautret et al conducted a study of 80 patients and reported that HCQ coadministration with AZI had a prompt reduction of viral load, with nearly 80-90% turned negative at Day 7-8 [12]. Another cohort study in France conducted by the same group enrolled 1061 new COVID-19 patients and treated with HCQ and AZI showed excellent clinical outcome for virological cure in 973 patients at day 10 (91.7%) with low mortality rate (0.75%) [15]. An Iraq study described 79 patients treated with a protocol that included HCQ and other antiviral drug combinations for different severity of COVID-19 had also showed a promising rate of recovery [14]. However, a small case series focused on severe-infected COVID-19 patients reported unfavorable outcome of viral elimination under the combination use of HCQ and AZI [13]. In present study, we could not exclude the influence of severity of COVID-19 in the outcome of combination use of HCQ and AZI due to the original data source, but this issue is important and worth further investigation.

The anti-inflammatory effect of HCQ on the immune system has been well recognized. The antiviral mechanisms of HCQ in treating COVID-19 patients has also been discovered, including increasing pH values of endosomes or lysosomes, stopping the glycosylation of angiotensin converting enzyme (ACE) 2 receptor, inhibiting antigen processing and following major histocompatibility complex (MHC)-II presentation to T cells and interrupting the interaction between cytosolic viral DNA/RNA and toll-like receptors (TLRs) [25, 26]. AZI, a member of the macrolide family, has antibacterial, anti-inflammatory, and antiviral properties [27, 28]. Although AZI has been found to block influenza virus internalization into host cells, its mechanism of action against SARS-CoV-2 is still uncertain [29]. Regarding the antiviral effects of using combination of HCQ and AZI, Andreani et al demonstrated that such combination showed a synergistic effect against SARS-CoV-2 when research was conducted *in vitro* [30].

In terms of safety issue of using AZI and HCQ, mild adverse effects such as rash and diarrhea have been described in trials and case series reports [7, 9, 11]. However, both AZI and HCQ are associated with QT prolongation during treatment, and combined use may potentiate this adverse effect [16, 31, 32]. In addition, our results showed that the incidence of mortality rate was higher in control group when comparing to the groups with HCQ treatment alone or combine with AZI (OR 2.54). We suggest that the sample size of individual study might have biased the outcome. While most of the included trials use small sample size in analysis, the study conducted by Magagnoli et al collected relatively large number of samples which did not exclude patients with contraindication of HCQ or significant comorbidities. Furthermore, the major population comprised in this study was non-Hispanic black men whose median age was over 65 years [10]. Numerous reports have indicated that the average age of COVID-19 patients was about 50 years, and the gender distribution was nearly equal [33–35]. However, the mortality risk for COVID-19 varies by age, older people might be related to multiple comorbidities and at a higher risk of COVID-19 -related hospitalization than young people [36–38]. Higher rates of COVID-19 -related hospitalization among the elder population and non-Hispanic black have also been found in the United States [39]. The mortality rate in different countries might be also associated with socio-economic and political factors [40]. Therefore, our present results of pooled analysis might be biased as we also included the demographic composition of patients, mentioned in the study conducted by Magagnoli et al.

Our study has demonstrated some substantial advantages. We performed a systematic and comprehensive search to locate published and unpublished papers through appropriate database. There were no potentially essential sources excluded with regard to language or methodological features. Consequently, the review process caused little publication-selection bias. Furthermore, we have used intention-to-treat methods and performed subgroup analysis with stable findings.

Despite the pertinent information demonstrated above, our systematic review has its own limitations. As far as the lack of compiling sufficient data with regard to virological cure as efficacy outcome is concerned, we tried to solve this problem by conducting a search for unpublished literature and non-English journals to increase the quantity of relevant integrated literature. Most studies which are published are mainly observational studies and case series with substantial methodology concerns such as no framework of exclusion criteria, and therefore susceptible to bias and confounding. Therefore, as there are limited related RCTs in the literature at present, only those were included in our study. In the lack of experimental random allocation to the intervention, the effects of the HCQ might be overestimated. Furthermore, the two single-center RCTs which were conducted in China had some limitations as it included small sample size, did not conduct an international trial, and only limited data was available regarding the viral load. In our present study, bias might be present since little information was provided on demographic data, comorbidity, severity of disease, and the adverse effects of HCQ or AZI such as QT prolongation. In addition, we have not performed a dose–response meta-analysis, due to the lack of original data. Finally, the limited number of available studies which were included, made it unworkable to evaluate the true effect of HCQ. Therefore, the present results should be interpreted cautiously, and further large, prospective RCTs needs to be conducted in a methodologically rigorous manner.

In conclusion, the present study has suggested that HCQ alone, or in combination with AZI for COVID-19 treatment showed beneficial outcome but also higher mortality. The combination of HCQ and AZI has demonstrated synergistic effects. In future, large, randomized, placebo-controlled trials with longer follow-up are needed to further confirm these findings.

## Data Availability

The authors confirm that the data supporting the findings of this study are available within the article and its supplementary materials.

## Acknowledgments

None.

## Conflict of interest

None declared.

## Funding

The authors disclosed receipt of the following financial support for the research, authorship, and/or publication of this article: This study was supported partly by Taipei 21Veterans General Hospital (V109EA-014, VGH-2017/2020 C and E project).

## Declaration of interest

The authors report no conflict of interest.

## Notes

### Competing Interest Statement

The authors have declared no competing interest.

## References

1. Steven S, Yen Ting L, Chonggang X, et al. High Contagiousness and Rapid Spread of Severe Acute Respiratory Syndrome Coronavirus 2. Emerg Infect Dis 2020; 26(7).

2. Worldometers. COVID-19 coronavirus pandemic. 2020; Available from: https://www.worldometers.info/coronavirus/.

3. Wang X, Xu W, Hu G, et al. SARS-CoV-2 infects T lymphocytes through its spike protein-mediated membrane fusion. Cell Mol Immunol 2020.

4. Colson P, Rolain JM, Lagier JC, et al. Chloroquine and hydroxychloroquine as available weapons to fight COVID-19. International journal of antimicrobial agents 2020; 55(4): 105932.

5. Yao X, Ye F, Zhang M, et al. In Vitro Antiviral Activity and Projection of Optimized Dosing Design of Hydroxychloroquine for the Treatment of Severe Acute Respiratory Syndrome Coronavirus 2 (SARS-CoV-2). Clinical infectious diseases: an official publication of the Infectious Diseases Society of America 2020.

6. Liu J, Cao R, Xu M, et al. Hydroxychloroquine, a less toxic derivative of chloroquine, is effective in inhibiting SARS-CoV-2 infection in vitro. Cell discovery 2020; 6: 16.

7. Chen J, Liu D, Liu L, et al. A pilot study of hydroxychloroquine in treatment of patients with common coronavirus disease-19 (COVID-19). J Zhejiang Univ (Med Sci) 2020; Published online March 6, 2020. doi:10.3785/j.issn.1008-9292.2020.03.03.

8. Gautret P, Lagier J, Parola P, et al. Hydroxychloroquine and azithromycin as a treatment of COVID-19: results of an open-label non-randomized clinical trial. International journal of antimicrobial agents 2020; Published online March 20, 2020. doi: 10.1016/j.ijantimicag.2020.105949:105949.

9. Chen Z, Hu H, Zhang Z, et al. Efficacy of hydroxychloroquine in patients with COVID-19: results of a randomized clinical trial. medRxiv 2020: preprint. [https://doi.org/10.1101/2020.03.22.20040758].

10. Magagnoli J, Narendran S, Pereira F, et al. Outcomes of hydroxychloroquine usage in United States veterans hospitalized with Covid-19. medRxiv 2020: preprint. [https://doi.org/10.1101/2020.04.16.20065920].

11. Mahevas M, Tran V, Roumier M, et al. No evidence of clinical efficacy of hydroxychloroquine in patients hospitalized for COVID-19 infection with oxygen requirement: results of a study using routinely collected data to emulate a target trial. medRxiv 2020: preprint. [https://doi.org/10.1101/2020.04.10.20060699].

12. Gautret P, Lagier J, Parola P, et al. Clinical and microbiological effect of a combination of hydroxychloroquine and azithromycin in 80 COVID-19 patients with at least a six-day follow up: A pilot observational study. Travel Med Infect Dis 2020: 101663.

13. Molina J, Delaugerre C, Le Goff J, et al. No Evidence of Rapid Antiviral Clearance or Clinical Benefit with the Combination of Hydroxychloroquine and Azithromycin in Patients with Severe COVID-19 Infection. Medecine et maladies infectieuses 2020.

14. Allawi J, Abbas H, Rasheed J, et al. The first 40-days experience and clinical outcomes in the management of coronavirus covid-19 crisis. Single center preliminary study. J Fac Med Bagdad [Internet] 2020; 61(3,4): Available from: http://iqjmc.uobaghdad.edu.iq/index.php/19JFacMedBaghdad36/article/view/1739.

15. Million M, Lagier J-C, Gautret P, et al. Full-length title: Early treatment of COVID-19 patients with hydroxychloroquine and azithromycin: A retrospective analysis of 1061 cases in Marseille, France. Travel Medicine and Infectious Disease 2020: 101738.

16. Chorin E, Dai M, Shulman E, et al. The QT Interval in Patients with SARS-CoV-2 Infection Treated with Hydroxychloroquine/Azithromycin. medRxiv 2020: preprint. [https://doi.org/10.1101/2020.04.02.20047050].

17. Gbinigie K, Frie K. Should chloroquine and hydroxychloroquine be used to treat COVID-19? A rapid review. BJGP Open 2020: bjgpopen20X101069.

18. Singh A, Singh A, Shaikh A, et al. Chloroquine and hydroxychloroquine in the treatment of COVID-19 with or without diabetes: A systematic search and a narrative review with a special reference to India and other developing countries. Diabetes & metabolic syndrome 2020; 14(3): 241–6.

19. Kapoor K, Kapoor A. Role of Chloroquine and Hydroxychloroquine in the Treatment of COVID-19 Infection- A Systematic Literature Review. medRxiv 2020: preprint. [https://doi.org/10.1101/2020.03.24.20042366].

20. Sarma P, Kaur H, Kumar H, et al. Virological and Clinical Cure in Covid-19 Patients Treated with Hydroxychloroquine: A Systematic Review and Meta-Analysis. Journal of medical virology 2020.

21. Shamshirian A, Hessami A, Heydari K, et al. Hydroxychloroquine Versus COVID-19: A Rapid Systematic Review and Meta-Analysis. medRxiv 2020: preprint. [https://doi.org/10.1101/2020.04.14.20065276].

22. FDA. FDA cautions against use of hydroxychloroquine or chloroquine for COVID-19 outside of the hospital setting or a clinical trial due to risk of heart rhythm problems. FDA Drug Safety Communication 2020: https://www.fda.gov/drugs/drug-safety-and-availability/fda-cautions-against-use-hydroxychloroquine-or-chloroquine-covid-19-outside-hospital-setting-or.

23. Tang W, Cao Z, Han M, et al. Hydroxychloroquine in patients with COVID-19: an open-label, randomized, controlled trial. medRxiv 2020: preprint. [https://doi.org/10.1101/2020.04.10.20060558].

24. Downs S, Black N. The feasibility of creating a checklist for the assessment of the methodological quality both of randomised and non-randomised studies of health care interventions. Journal of epidemiology and community health 1998; 52(6): 377–84.

25. Alia E, Grant-Kels JM. Does Hydroxychloroquine Combat COVID-19? A Timeline of Evidence. J Am Acad Dermatol 2020.

26. Sinha N, Balayla G. Hydroxychloroquine and covid-19. Postgrad Med J 2020.

27. Schogler A, Kopf BS, Edwards MR, et al. Novel antiviral properties of azithromycin in cystic fibrosis airway epithelial cells. The European respiratory journal 2015; 45(2): 428–39.

28. Gielen V, Johnston SL, Edwards MR. Azithromycin induces anti-viral responses in bronchial epithelial cells. The European respiratory journal 2010; 36(3): 646–54.

29. Ohe M, Shida H, Jodo S, et al. Macrolide treatment for COVID-19: Will this be the way forward? Bioscience trends 2020.

30. Andreani J, Le Bideau M, Duflot I, et al. In vitro testing of combined hydroxychloroquine and azithromycin on SARS-CoV-2 shows synergistic effect. Microbial Pathogenesis 2020; 145: 104228.

31. Mercuro NJ, Yen CF, Shim DJ, et al. Risk of QT Interval Prolongation Associated With Use of Hydroxychloroquine With or Without Concomitant Azithromycin Among Hospitalized Patients Testing Positive for Coronavirus Disease 2019 (COVID-19). JAMA cardiology 2020.

32. Bessiere F, Roccia H, Deliniere A, et al. Assessment of QT Intervals in a Case Series of Patients With Coronavirus Disease 2019 (COVID-19) Infection Treated With Hydroxychloroquine Alone or in Combination With Azithromycin in an Intensive Care Unit. JAMA cardiology 2020.

33. Tang C, Zhang K, Wang W, et al. Clinical Characteristics of 20,662 Patients with COVID-19 in mainland China: A Systemic Review and Meta-analysis. medRxiv 2020: preprint. [https://doi.org/10.1101/2020.04.18.20070565].

34. Wu J, Liu J, Zhao X, et al. Clinical Characteristics of Imported Cases of Coronavirus Disease 2019 (COVID-19) in Jiangsu Province: A Multicenter Descriptive Study. Clinical Infectious Diseases 2020.

35. Qian K, Deng Y, Tai Y, et al. Clinical Characteristics of 2019 Novel Infected Coronavirus Pneumonia A Systemic Review and Meta-analysis. medRxiv 2020: preprint. [https://doi.org/10.1101/2020.02.14.20021535].

36. Zhai P, Ding Y, Wu X, et al. The epidemiology, diagnosis and treatment of COVID-19. International journal of antimicrobial agents 2020: 105955.

37. Novel Coronavirus Pneumonia Emergency Response Epidemiology Team. The epidemiological characteristics of an outbreak of 2019 novel coronavirus diseases (COVID-19) in China. Zhonghua liu xing bing xue za zhi 2020; 41(2): 145–51.

38. Zhou F, Yu T, Du R, et al. Clinical course and risk factors for mortality of adult inpatients with COVID-19 in Wuhan, China: a retrospective cohort study. Lancet 2020; 395(10229): 1054–62.

39. Garg S, Kim L, Whitaker M. Hospitalization Rates and Characteristics of Patients Hospitalized with Laboratory-Confirmed Coronavirus Disease 2019 — COVID-NET, 14 States, March 1–30, 2020. MMWR Morb Mortal Wkly Rep 2020; (69): 458-64.

40. Sorci G, Faivre B, Morand S. Why does COVID-19 case fatality rate vary among countries? medRxiv 2020: preprint. [https://doi.org/10.1101/2020.04.17.20069393].

